# Deep Learning in Myocarditis: A Novel Approach to Severity Assessment

**DOI:** 10.1101/2025.08.01.25332620

**Authors:** Makoto Nishimori, Tomoyuki Otani, Yasuhide Asaumi, Keiko Ogo, Yoshihiko Ikeda, Kisaki Amemiya, Teruo Noguchi, Chisato Izumi, Masakazu Shinohara, Kinta Hatakeyama, Kunihiro Nishimura

**Affiliations:** Division of Molecular Epidemiology, Kobe University Graduate School of Medicine, Kobe, Japan; Department of Preventive Medicine and Epidemiology, National Cerebral and Cardiovascular Center, Osaka, Japan; Department of Pathology, National Cerebral and Cardiovascular Center, Osaka, Japan; Department of Cardiology, National Cerebral and Cardiovascular Center, Osaka, Japan

**Keywords:** deep learning, myocarditis, pathology

## Abstract

**Background:** Myocarditis is a life-threatening disease with significant hemodynamic risks during the acute phase. Although histopathological examination of myocardial biopsy specimens remains the gold standard for diagnosis, there is no established method for objectively quantifying cardiomyocyte damage. We aimed to develop an AI model to evaluate clinical myocarditis severity using comprehensive pathology data.

**Methods:** We retrospectively analyzed 314 patients (1076 samples) who underwent myocardial biopsy from 2002 to 2021 at the National Cerebrovascular Center. Among these patients, 158 were diagnosed with myocarditis based on the Dallas criteria. A Multiple Instance Learning (MIL) model served as a pre-trained classifier to detect myocarditis across whole-slide images. We then constructed two clinical severity-prediction models: (1) a logistic regression model (Model 1) using the density of inflammatory cells per unit area, and (2) a Transformer-based model (Model 2), which processed the top-ranked patches identified by the MIL model to predict clinical severe outcomes.

**Results:** Model 1 achieved an AUROC of 0.809, indicating a robust association between inflammatory cell density and severe myocarditis. In contrast, Model 2, the Transformer-based approach, yielded an AUROC of 0.993 and demonstrated higher accuracy and precision for severity prediction. Attention score visualizations showed that Model 2 captured both inflammatory cell infiltration and additional morphological features. These findings suggest that combining MIL with Transformer architectures enables more comprehensive identification of key histological markers associated with clinical severe disease.

**Conclusions:** Our results highlight that a Transformer-based AI model analyzing whole-slide pathology images can accurately assess clinical myocarditis severity. Moreover, simply quantifying the extent of inflammatory cell infiltration also correlates strongly with clinical outcomes. These methods offer a promising avenue for improving diagnostic precision, guiding treatment decisions, and ultimately enhancing patient management. Future prospective studies are warranted to validate these models in broader clinical settings and facilitate their integration into routine pathological workflows.

**What is new?:**

– This is the first study to apply an AI model for the diagnosis and severity assessment of myocarditis.
– New evidence shows that inflammatory cell infiltration is related to the severity of myocarditis.
– Using information from the entire tissue, not just inflammatory cells, allows for a more accurate assessment of myocarditis severity.

**What are the clinical implications?:**

– The use of the AI model allows for an unprecedented histological evaluation of myocarditis severity, which can enhance early diagnosis and intervention strategies.
– Rapid and precise assessments of myocarditis severity by the AI model can support clinicians in making timely and appropriate treatment decisions, potentially improving patient outcomes.
– The incorporation of this AI model into clinical practice may streamline diagnostic workflows and optimize the allocation of medical resources, enhancing overall patient care.

## Introduction

Myocarditis is a very serious and high-risk disease carrying the risk of circulatory fluctuations during the acute phase ^1^ ^2^ ^3^. Although clinical assessment typically involves symptoms, laboratory findings, and electrocardiograms, the definitive diagnosis relies on histopathological evaluation of myocardial biopsy^4^. Early assessment of clinical disease severity is also essential for guiding treatment and estimating prognosis^5^.

Although a study evaluating the clinical severity of myocarditis from histopathology have been reported^6^, an accurate index for evaluation has not yet been established. A factor that histologically defines the severity of myocarditis is the degree of myocardial cell damage. However, there is no method for morphologically objective assessment of cardiomyocyte damage, such as coarsening of the myocyte cytoplasm, irregular shape, or disorganization of the arrangement of myocytes. Therefore, we aimed to develop an artificial intelligence (AI)-based model capable of detecting myocardial tissue features indicative of clinical disease severity.

In previous studies on pathology AI models, various deep learning models have been developed for analyzing pathological tissues, including myocarditis ^7^ ^8^ ^9^ ^10^. While AI models for diagnosing myocarditis have been reported, no models specifically designed to assess the clinical severity of myocarditis have been developed. Achieving this goal requires a model capable of comprehensively interpreting whole-slide pathology images (WSIs). However, these images tend to be extremely large, making direct input into the model inefficient in terms of both computational resources and cost. Multiple Instance Learning (MIL) ^11^ provides a potential solution by dividing slides into smaller regions for training. In addition, MIL does not require detailed pixel-level annotations, instead using slide-level labels, which is particularly advantageous for pathology applications ^12^. Transformer architectures ^13^—now widely used as the backbone of large language models—have recently demonstrated high learning efficiency across multiple domains. To date, there are no reports of combining MIL with Transformer-based methods for myocardial pathology. We anticipate that this combination could significantly enhance performance in analyzing myocardial tissue. Therefore, the objective of this study is to develop a high-accuracy AI model capable of comprehensively interpreting WSIs to evaluate the clinical severity of myocarditis, ultimately aiming for clinical applicability.

## Methods

### Study Design, Data Sources, and Patient Population

This retrospective cohort study included 314 patients (1076 samples) who underwent myocardial biopsy at the National Cerebrovascular Center (NCVC) from January 2002 to December 2021. The cohort comprised 158 myocarditis and 160 non-myocarditis patients, the latter monitored post-heart transplant showing no signs of acute cellular rejection (ACR Grade 0). Our clinical study adhered to relevant guidelines, approved by the Ethics Committee of the NCVC (Approval No. 12232021-1-1), and was conducted in accordance with the Declaration of Helsinki.

### Diagnostic and Severity Criteria for Myocarditis

Diagnosis of myocarditis was determined by three cardiac pathologists based on the comprehensive Dallas criteria ^4^. The severity of myocarditis was defined by short-term in-hospital outcomes, including mortality, cardiogenic shock, or the need for mechanical support —namely intra-aortic balloon pumping, an Impella device, extracorporeal membrane oxygenation, or a ventricular assist device.

### Whole Slide Image Data

Hematoxylin and eosin-stained slides were digitized using a Hamamatsu NanoZoomer S210 C13239 series at a Source lens magnification of 40x, capturing detailed virtual slide images.

### Data Preprocessing

We used the openslide (Python) library ^14^ to read and process each ndpi file. To accommodate the large size of whole-slide images (WSIs) while retaining detailed histological features, we segmented each slide into 256×256-pixel patches. These dimensions were chosen based on preliminary experiments and prior pathology studies, which suggested a balance between sufficient tissue context and computational feasibility ^15^. Patches containing less than 50% tissue (based on simple thresholding of non-background pixels) were excluded to reduce noise from empty or artifact-laden regions. Data augmentation (e.g., random horizontal/vertical flips, rotations, and slight color jitter) was applied to mitigate overfitting and improve the model’s robustness to variations in staining and tissue orientation. ^16^

### Pretraining Model

To efficiently extract features indicative of myocarditis, a pretraining model was developed using all cases. The cases were randomly divided into training (70%), validation (15%), and test datasets (15%). A MIL model was employed, which does not require pixel-level annotations but uses slide-level labels for presence or absence of myocarditis.

### Model Development and Training

In this study, we developed two distinct approaches for severity prediction, each leveraging a different methodology. We then evaluated and compared their prognostic performance to determine which approach could more accurately predict clinical outcomes.

### Model1 (Inflammatory Cell Detection and Logistic Regression)

We focused on quantifying inflammatory infiltration as an initial, straightforward indicator of disease severity. First, two board-certified pathologists annotated lymphocytes on randomly selected patches. Lymphocytes were defined as mononuclear inflammatory cells with round nuclei, dense chromatin, and scant cytoplasm. All annotations were based on the consensus of the two pathologists. Using these annotations, we trained an object detection model based on YOLOv8 model ^17^ to count inflammatory cells per unit tissue area. A total of 1,000 annotated images were used; these were split into training (70%), validation (15%), and test (15%) sets. Default YOLOv8 hyperparameters were employed. We evaluated the object detection performance using metrics such as mean average precision on the validation set. The resulting feature for each 256×256 patch was the average lymphocytes count, computed by summing YOLOv8 detections and normalizing by tissue area.

Finally, we applied a univariate logistic regression model, using the lymphocytes density as the sole independent variable to predict disease severity. This univariate approach enabled us to isolate the contribution of lymphocytes infiltration, with future plans to incorporate additional factors for more comprehensive modeling.

### Model2 (Transformer-Based Severity Classification)

Model 2 employed a Transformer-based approach to classify disease severity. We began by applying a pretrained MIL model to estimate the likelihood of myocarditis for each patch. To reduce computational demands and focus on the most informative regions, we selected the top N patches showing the highest probabilities of myocarditis. These patches were then encoded into embedding vectors and processed by a Transformer encoder along with a Class Token. The final classification layer—a multi-layer perceptron applied to the Class Token output—performed a binary classification (severe vs. non-severe) via a sigmoid function. All major hyperparameters, including the Transformer architecture (e.g., number of layers, attention heads), the learning rate, and batch size, were optimized using Bayesian optimization ^18^. Training was conducted in parallel on two workstations, each equipped with an NVIDIA GeForce RTX 4090 GPU, to expedite processing. Validation set performance guided hyperparameter tuning, ensuring the model converged effectively on the classification task.

### Decision Rationale Visualization

To elucidate the contributions of different regions to the severity diagnosis, we visualized the attention scores assigned to each patch by the Class Token in the Transformer model ^13^, indicating key areas impacting the severity assessment (Figure 5).

### Statistics

Model performance was assessed through precision, recall, F1-score, area under the receiver operating characteristic curve (AUROC), and area under the precision-recall curve (AUPRC). Overall accuracy was also evaluated, with 95% confidence intervals calculated using the bootstrap method ^19^. P-values were determined using two-sided significance tests.

### Software

The analysis was conducted using Python 3.8, with PyTorch 1.8 ^20^ for deep learning tasks and scikit-learn for statistical calculations.

## Results

### Patients Characteristics

The study period spanned from January 2002 to December 2021 at the NCVC. A total of 145 patients diagnosed with myocarditis, including children, were eligible for inclusion. These patients had undergone myocardial biopsy and were confirmed to have myocarditis. No specific exclusion criteria were set.

The average age of these patients was 45 years (±21.46 years), with 59% being male. Among the total cohort, 88 patients (60.7%) were classified as severe cases. The average peak creatine kinase-MB values were 135.43 for the entire cohort, 166.83 for the severe group, and 54.99 for the non-severe group. Detailed information on other laboratory data at the onset for each group is provided in Table 1.

**Table 1:**
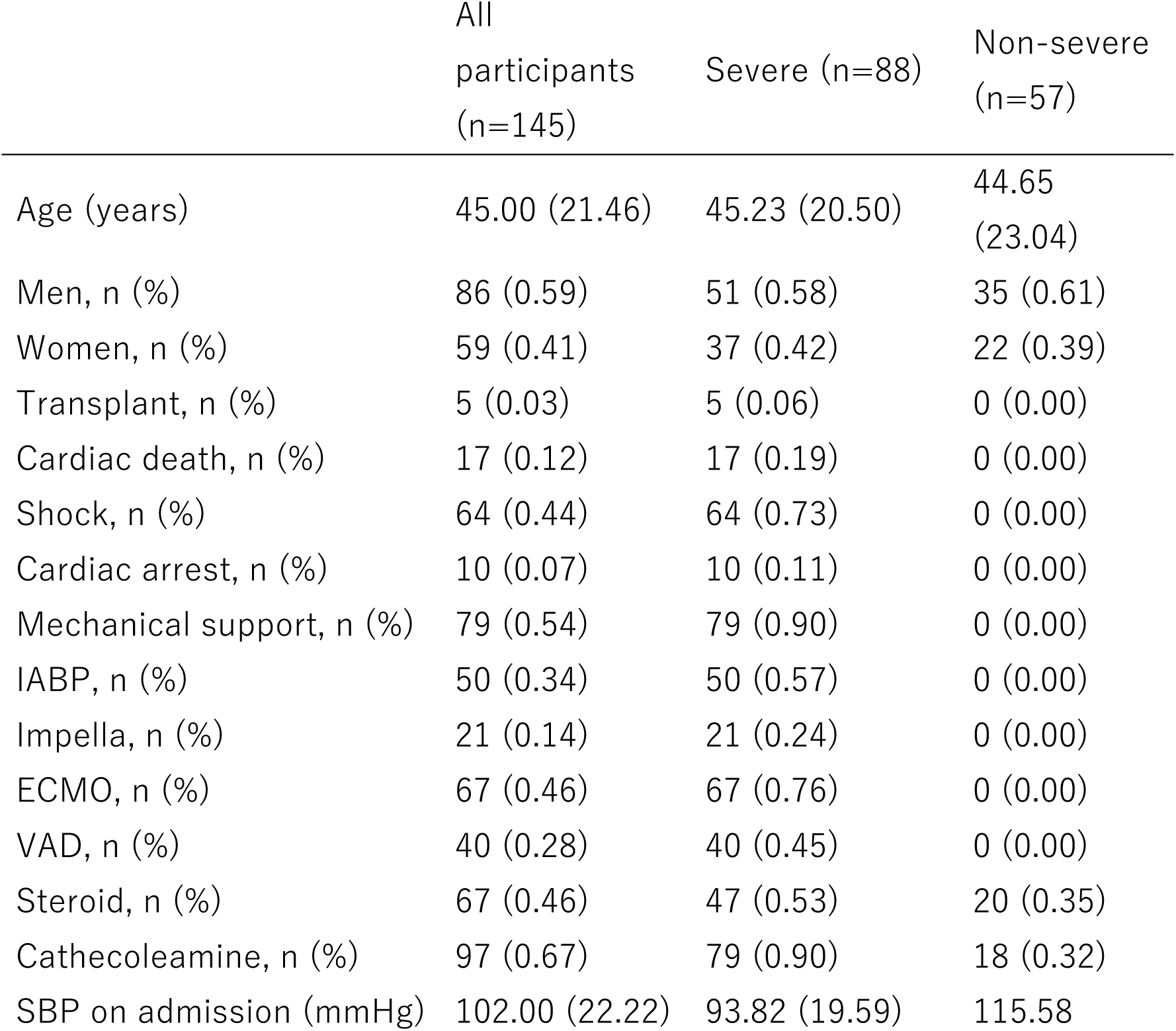
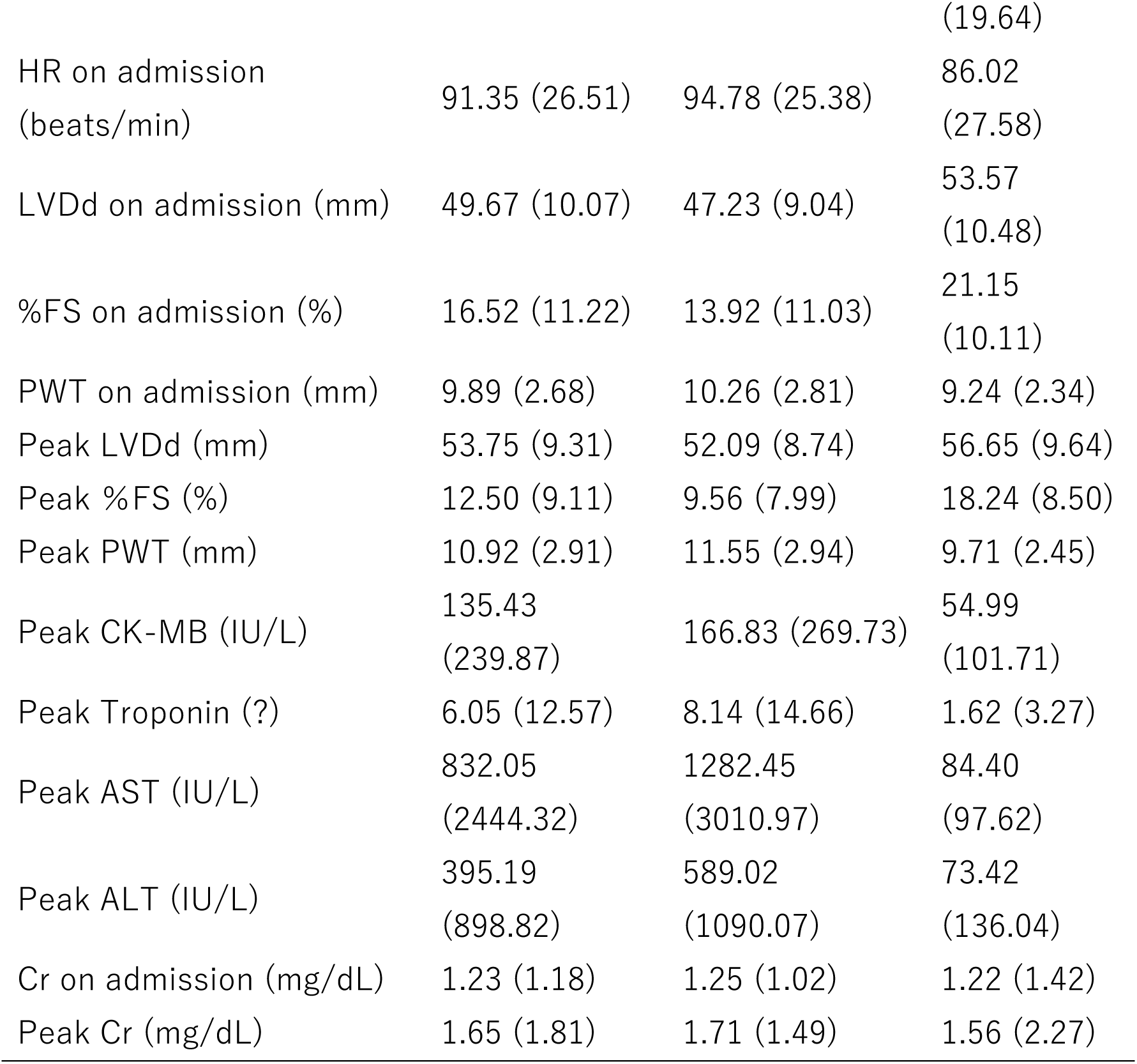
Patient characteristics.

### Pre-training with Multiple Instance Learning Model

A pre-training model was developed using MIL on a dataset of 314 patients with 1076 myocardial pathology samples, including myocarditis patients, to diagnose myocarditis (Table 2). The results from the training dataset showed an accuracy of 0.751 and an AUCROC of 0.717. In the validation dataset, the model achieved an accuracy of 0.749 and an AUCROC of 0.712. Visualization of the diagnostic rationale in the inference phase was performed, with heatmaps illustrating the areas diagnosed as myocarditis shown in Supplemental Figure 1. The heatmaps indicate higher values in regions where inflammatory cells are more concentrated.

**Table 2:**
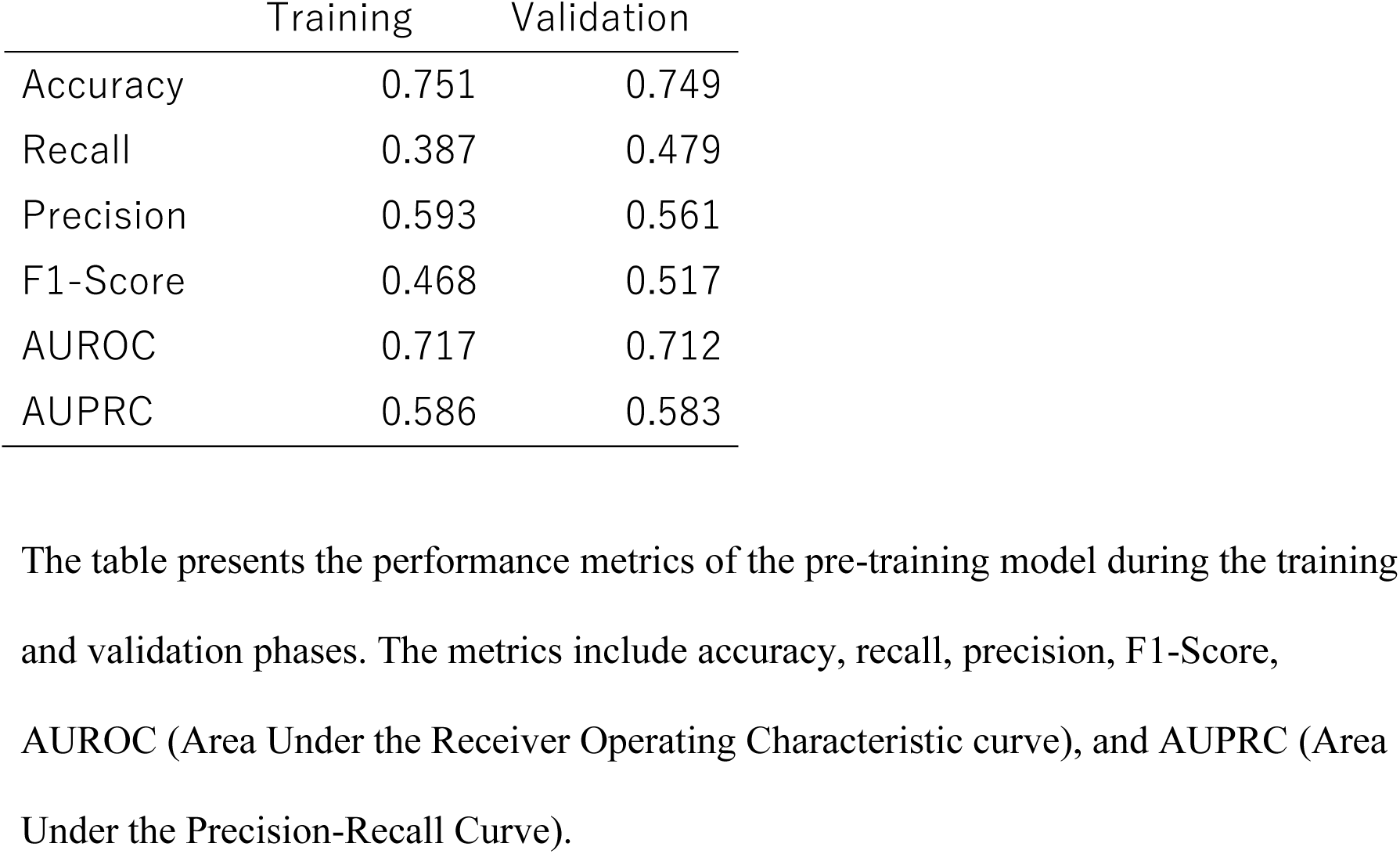
Results of pre-training model.

### Model 1: Predicting Myocarditis Severity Using a Deep Learning Model Based on Inflammatory Cell Infiltration

An object detection model (YOLOv8) was pre-trained to detect lymphocytes cells, based on annotations by two myocardial pathology specialists. This model was used to automatically detect the number of lymphocytes, and a logistic regression model was constructed using the number of lymphocytes per unit area as a variable to evaluate clinical myocarditis severity. The results of inflammatory cell detection by the Object Detection Model are shown in Supplemental Figure 1. Histogram plots of lymphocytes count per 256×256 pixel patch showed that clinical severe myocarditis cases had relatively higher lymphocytes counts per patch, while non-severe cases had lower counts (Figure 3-A). When converted to the number of lymphocytes per unit area (1 mm²), high values were observed in clinical severe cases, but most overlapped with non-severe cases (Figure 3-B). The logistic regression model using the number of lymphocytes per unit area as a variable resulted in an AUROC of 0.809 (Figure 4-A). Histograms of the predicted values for clinical severe and non-severe cases were also shown (Figure 4-B).

**Figure 1:**
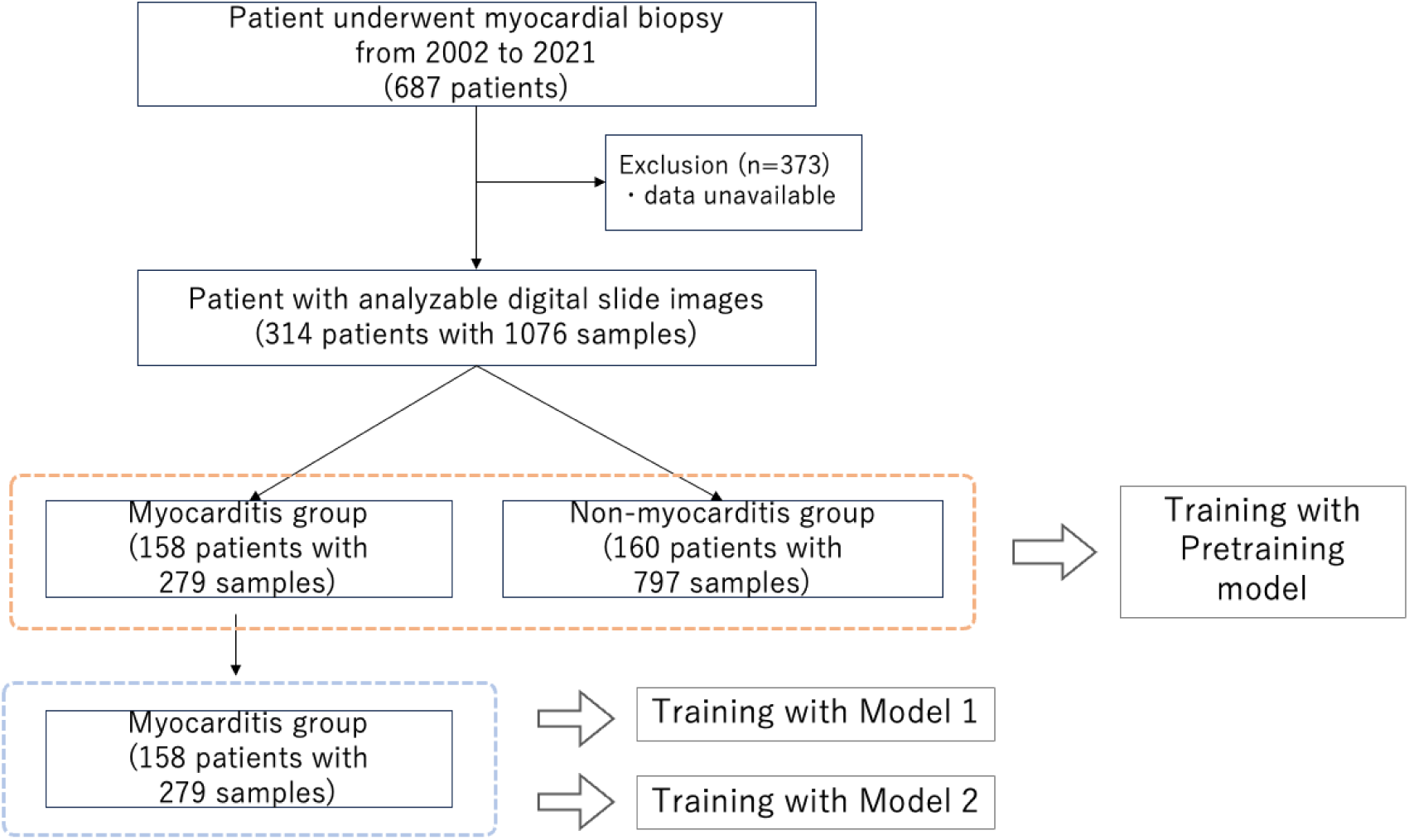
Study Cohort and Analysis Workflow. The flowchart shows the selection and analysis process for patients undergoing myocardial biopsy from 2002 to 2021. Out of 687 patients, 373 were excluded, leaving 314 patients with 1076 samples. These were divided into myocarditis (158 patients, 279 samples) and non-myocarditis (160 patients, 797 samples) groups. The myocarditis group was used for training two models after pre-training with a MIL model.

**Figure 2:**
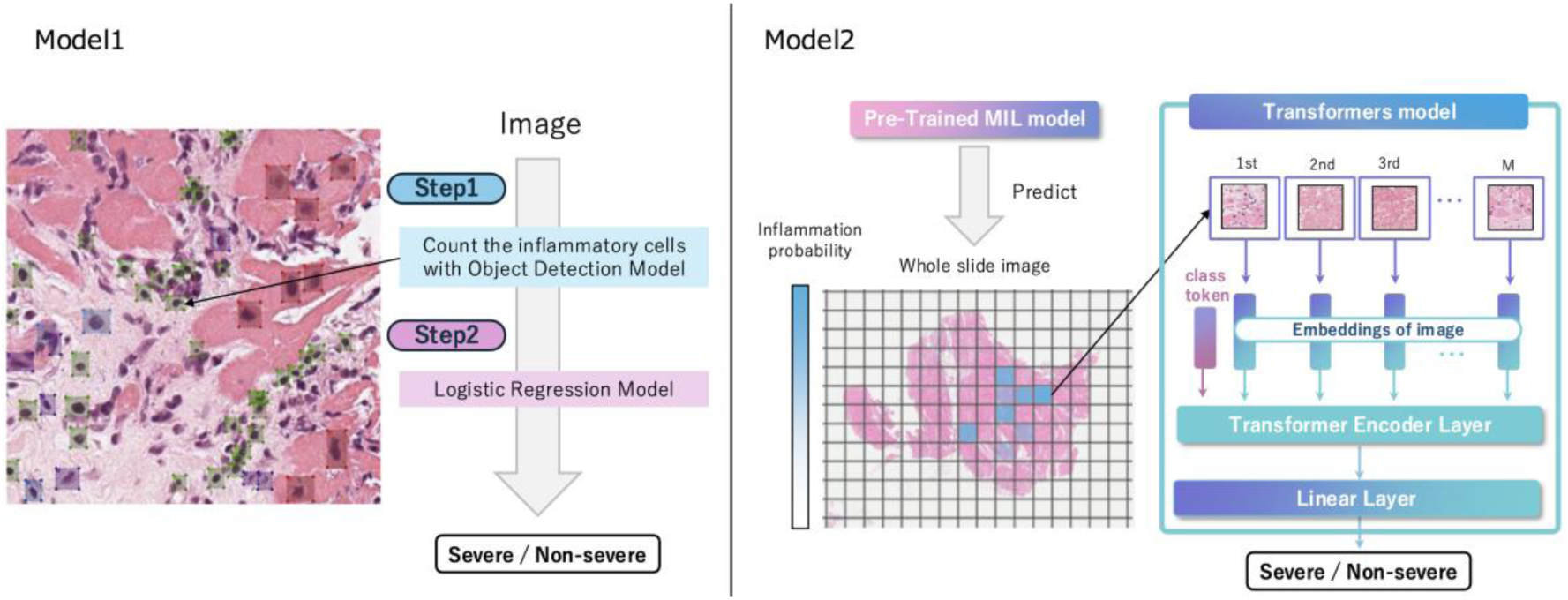
AI Models for Severity Classification. The figure shows two AI models used to classify myocarditis severity. Model 1: An object detection AI model counts inflammatory cells, followed by a logistic model to classify severity as severe or non-severe. Model 2: A trained MIL model predicts myocarditis probability. The MIL features are processed through a Transformer model to provide the final diagnosis of severe or non-severe.

**Figure 3:**
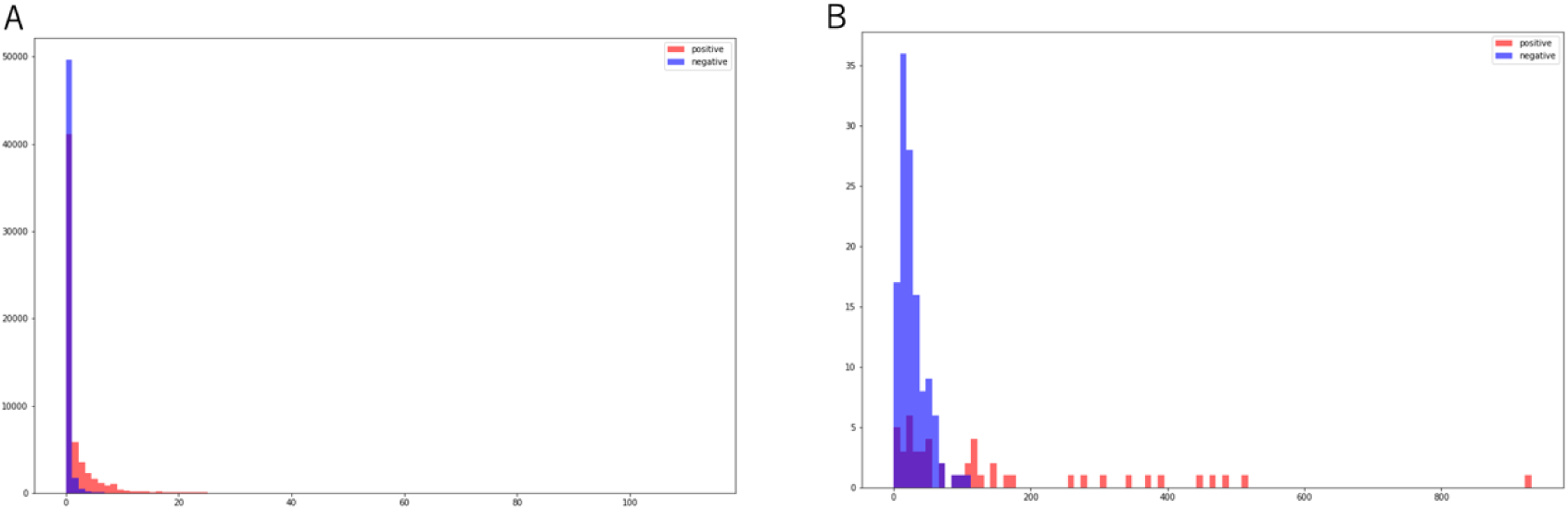
Distribution Plot of Lymphocytes Cells Detection Model. (A) The distribution of lymphocytes counts per patch, comparing severe and non-severe samples. The x-axis represents the number of lymphocytes per patch, and the y-axis represents the frequency of these counts. (B) The distribution of lymphocytes counts per 1mm² for each patient, comparing severe and non-severe samples. The x-axis represents the number of lymphocytes per 1mm², and the y-axis represents the frequency of these counts.

**Figure 4:**
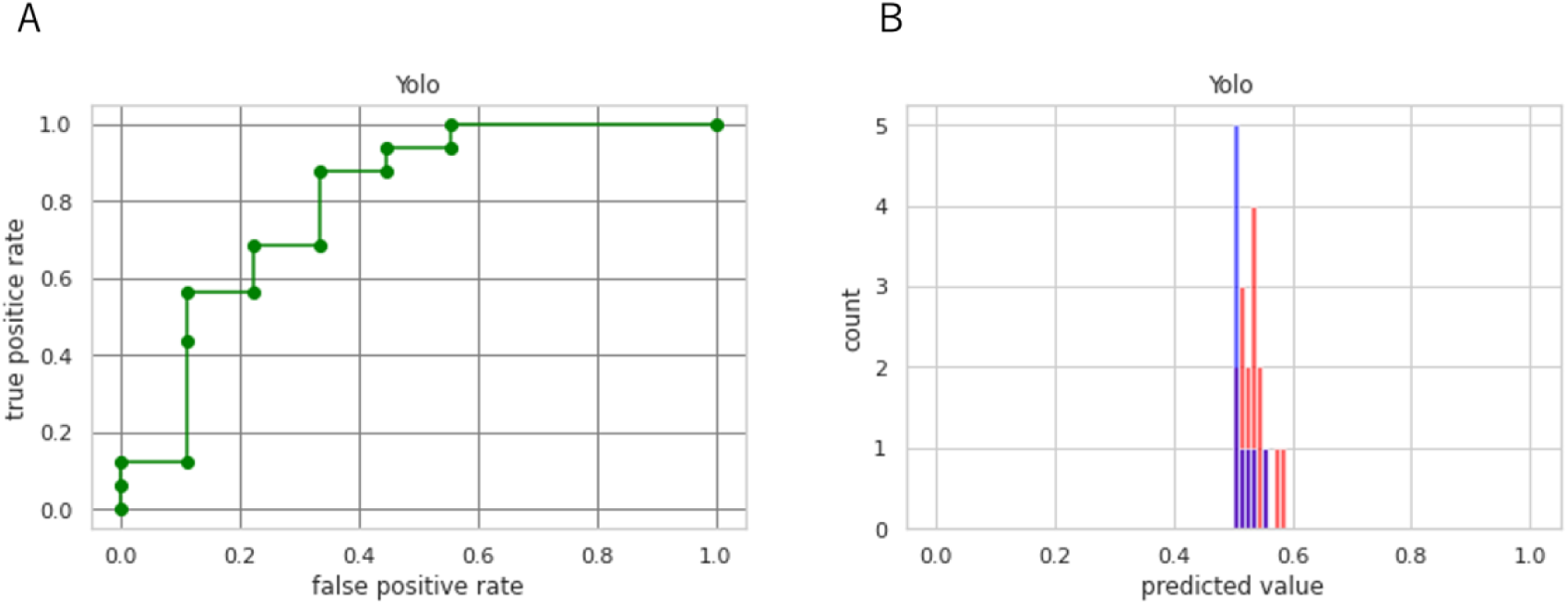
Results of Model 1. (A) Receiver Operating Characteristic (ROC) curve for Model 1. The x-axis represents the false positive rate, and the y-axis represents the true positive rate. Each point on the curve represents a different threshold value used to discriminate between classes. (B) Histogram of predicted values for Model 1. The x-axis represents the predicted value, and the y-axis represents the count of predictions. The red bars indicate positive samples, and the blue bars indicate negative samples.

### Model 2: Predicting Myocarditis Severity Using a Transformer-Based Model

The pre-trained MIL model was fixed, and for each patch, the MIL model inferred the presence of myocarditis. The top N candidate patches were then input into a Transformer-based model to predict clinical myocarditis severity. The results showed an AUROC of 0.993 (0.952 - 1.000), an accuracy of 0.64 (0.480 - 0.800), a precision of 0.64 (0.480 - 0.800), and a recall of 1 (1 - 1) (Supplemental Table1). ROC curves and histograms of predicted values are shown in Figures 5-A and 5-B.

**Figure 5:**
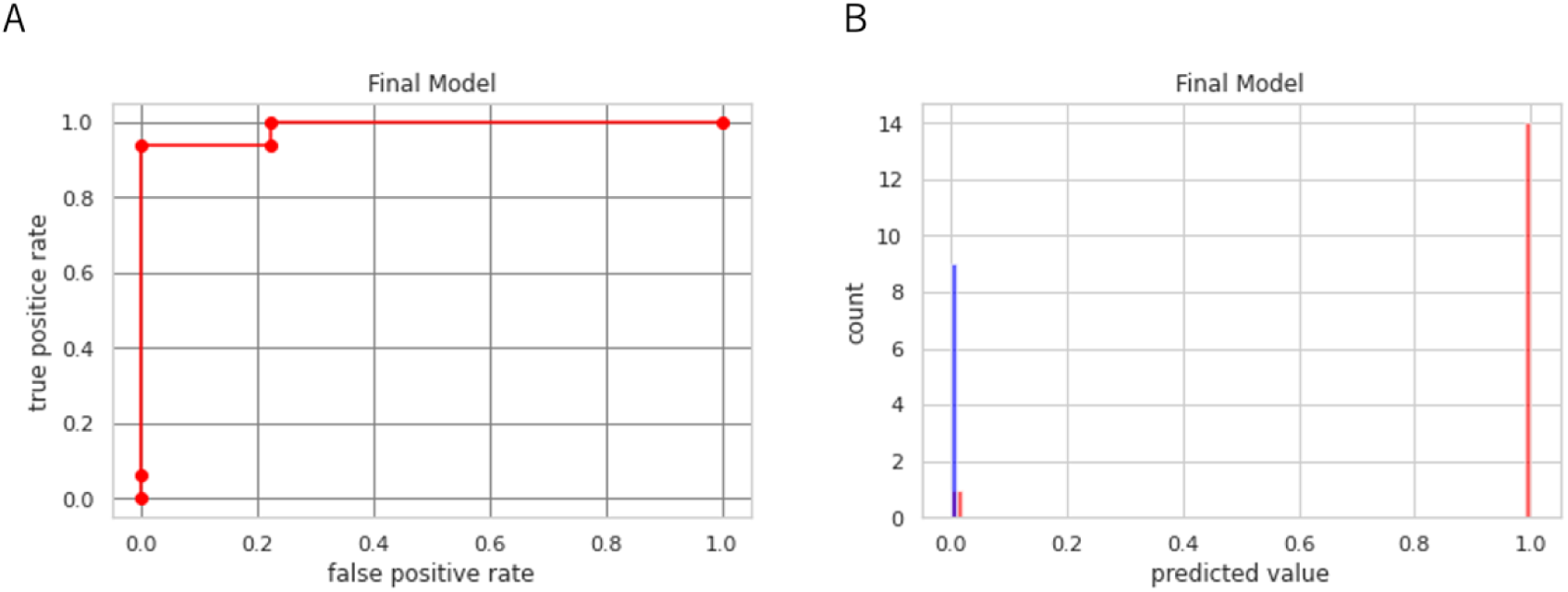
Results of Model 2. (A) Receiver Operating Characteristic (ROC) curve for Model 2. The x-axis represents the false positive rate, and the y-axis represents the true positive rate. Each point on the curve represents a different threshold value used to discriminate between classes. (B) Histogram of predicted values for Model 2. The x-axis represents the predicted value, and the y-axis represents the count of predictions. The red bars indicate positive samples, and the blue bars indicate negative samples.

### Comparison of Models for Predicting Myocarditis Severity

The metrics differences between Model 1 and Model 2 for predicting clinical severe myocarditis were evaluated using the bootstrap method. The Transformer-based Model 2 showed significantly higher accuracy, precision, and AUROC compared to Model 1.

### Visualization of Rationale for Severity Prediction

To visualize the rationale for clinical severe myocarditis prediction with Model 2, the attention scores of each input patch were illustrated for representative cases (Figure 6).

**Figure 6:**
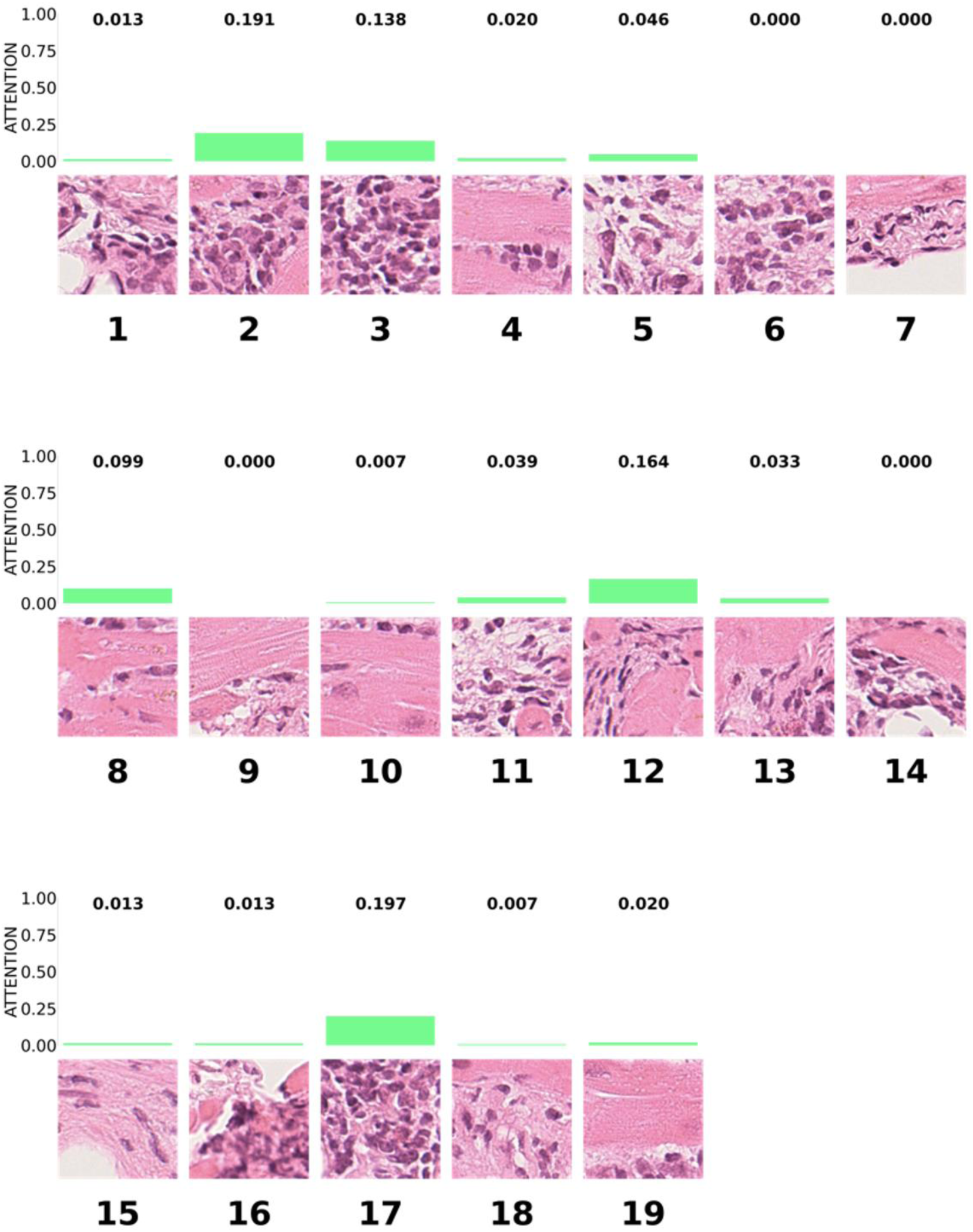
Attention Scores for Severe Myocarditis. The figure displays attention scores assigned to histological images of severe myocarditis by the model. Each image is associated with an attention score, indicating the model’s focus on different regions of the images. The green bars above each image represent the attention score, with higher bars indicating greater attention by the model.

## Discussion

In this study, we developed two AI models to evaluate the severity of myocarditis. Model 1, based on lymphocytes density, achieved an AUROC of 0.809, providing novel evidence that lymphocytes count correlate strongly with disease severity. In contrast, Model 2, which uses a Transformer-based approach to analyze whole-slide pathology images, attained a higher AUROC of 0.993. These findings suggest that a deep learning model capable of integrating diverse morphological features throughout the tissue may outperform a simpler model focused solely on the extent of inflammatory cell infiltration.

The clinical significance of Model 1 lies in its demonstration that inflammatory cell density is closely linked to disease severity. While immunological mechanisms have long been implicated in myocarditis ^21^, research quantifying inflammatory infiltrates as a direct prognostic marker remains sparse. Model 2, employing the Transformer architecture, captures subtle morphological and spatial patterns beyond mere cell counts. Unlike recurrent neural network (RNN) ^22^-based methods, Transformers incorporate broad contextual information, yielding more comprehensive pathological insights and higher predictive accuracy.

These findings have potential relevance for risk stratification and therapeutic decision-making in clinical practice. For example, quantification of inflammatory cell density could aid in the early identification of patients who may benefit from intensified immunosuppressive therapy or mechanical circulatory support. Although weakly supervised learning and RNN-based whole-slide analysis have been explored in other fields (e.g., oncology) ^15^, there is a scarcity of evidence demonstrating performance improvements via Transformers specifically in myocarditis. Our study thus contributes new data to this field and highlights the importance of considering histological features that extend beyond inflammatory cell infiltration.

### Limitations

Several limitations must be acknowledged. First, this is a retrospective analysis and therefore cannot establish causality. Additionally, the applicability of these models to diverse patient populations or specific myocarditis subtypes (such as giant cell myocarditis) remains insufficiently explored. Furthermore, factors including sample size and inter-institutional variability may affect the generalizability of our results.

## Conclusion

In conclusion, our results indicate that a Transformer-based deep learning approach offers more accurate severity prediction than a method reliant solely on inflammatory cell counts. At the same time, quantifying inflammatory infiltrates emerges as a potentially new and clinically relevant marker of disease severity. Moving forward, prospective and multi-center studies are needed to validate these models and integrate them into routine clinical workflows. By enabling more timely and precise therapeutic interventions, these AI-based models could ultimately improve patient outcomes and optimize healthcare resource allocation in the management of myocarditis.

## Abbreviations

AI: Artificial Intelligence
MIL: Multiple Instance Learning
WSIs: Whole-Slide pathology Images
AUROC: Area Under the Receiver Operating Characteristic curve
AUPRC: Area Under the Precision-Recall Curve
RNN: Recurrent Neural Network

## Data Availability

De-identified data are available on reasonable request to the corresponding author.

## References

1. Lorusso R, Centofanti P, Gelsomino S, Barili F, Di Mauro M, Orlando P, Botta L, Milazzo F, Actis Dato G, Casabona R, et al. Venoarterial Extracorporeal Membrane Oxygenation for Acute Fulminant Myocarditis in Adult Patients: A 5-Year Multi-Institutional Experience. Ann Thorac Surg. 2016;101:919–926. doi: 10.1016/j.athoracsur.2015.08.014

2. McCarthy RE, 3rd, Boehmer JP, Hruban RH, Hutchins GM, Kasper EK, Hare JM, Baughman KL. Long-term outcome of fulminant myocarditis as compared with acute (nonfulminant) myocarditis. N Engl J Med. 2000;342:690–695. doi: 10.1056/NEJM200003093421003

3. Gupta S, Markham DW, Drazner MH, Mammen PP. Fulminant myocarditis. Nat Clin Pract Cardiovasc Med. 2008;5:693–706. doi: 10.1038/ncpcardio1331

4. Aretz HT, Billingham ME, Edwards WD, Factor SM, Fallon JT, Fenoglio JJ, Jr., Olsen EG, Schoen FJ. Myocarditis. A histopathologic definition and classification. Am J Cardiovasc Pathol. 1987;1:3–14.

5. Ammirati E, Frigerio M, Adler ED, Basso C, Birnie DH, Brambatti M, Friedrich MG, Klingel K, Lehtonen J, Moslehi JJ, et al. Management of Acute Myocarditis and Chronic Inflammatory Cardiomyopathy: An Expert Consensus Document. Circ Heart Fail. 2020;13:e007405. doi: 10.1161/CIRCHEARTFAILURE.120.007405

6. Kanaoka K, Onoue K, Terasaki S, Nakano T, Nakai M, Sumita Y, Hatakeyama K, Terasaki F, Kawakami R, Iwanaga Y, et al. Features and Outcomes of Histologically Proven Myocarditis With Fulminant Presentation. Circulation. 2022;146:1425–1433. doi: 10.1161/CIRCULATIONAHA.121.058869

7. Song Z, Zou S, Zhou W, Huang Y, Shao L, Yuan J, Gou X, Jin W, Wang Z, Chen X, et al. Clinically applicable histopathological diagnosis system for gastric cancer detection using deep learning. Nat Commun. 2020;11:4294. doi: 10.1038/s41467-020-18147-8

8. Yang H, Chen L, Cheng Z, Yang M, Wang J, Lin C, Wang Y, Huang L, Chen Y, Peng S, et al. Deep learning-based six-type classifier for lung cancer and mimics from histopathological whole slide images: a retrospective study. BMC Med. 2021;19:80. doi: 10.1186/s12916-021-01953-2

9. Jiang B, Bao L, He S, Chen X, Jin Z, Ye Y. Deep learning applications in breast cancer histopathological imaging: diagnosis, treatment, and prognosis. Breast Cancer Res. 2024;26:137. doi: 10.1186/s13058-024-01895-6

10. Moravvej SV, Alizadehsani R, Khanam S, Sobhaninia Z, Shoeibi A, Khozeimeh F, Sani ZA, Tan RS, Khosravi A, Nahavandi S, et al. RLMD-PA: A Reinforcement Learning-Based Myocarditis Diagnosis Combined with a Population-Based Algorithm for Pretraining Weights. Contrast Media Mol Imaging. 2022;2022:8733632. doi: 10.1155/2022/8733632

11. Andrews S, Tsochantaridis I, Hofmann T. Support vector machines for multiple-instance learning. Advances in neural information processing systems. 2002;15.

12. Gadermayr M, Tschuchnig M. Multiple instance learning for digital pathology: A review of the state-of-the-art, limitations & future potential. Comput Med Imaging Graph. 2024;112:102337. doi: 10.1016/j.compmedimag.2024.102337

13. Vaswani A. Attention is all you need. Advances in Neural Information Processing Systems. 2017.

14. Goode A, Gilbert B, Harkes J, Jukic D, Satyanarayanan M. OpenSlide: A vendor-neutral software foundation for digital pathology. J Pathol Inform. 2013;4:27. doi: 10.4103/2153-3539.119005

15. Campanella G, Hanna MG, Geneslaw L, Miraflor A, Werneck Krauss Silva V, Busam KJ, Brogi E, Reuter VE, Klimstra DS, Fuchs TJ. Clinical-grade computational pathology using weakly supervised deep learning on whole slide images. Nat Med. 2019;25:1301–1309. doi: 10.1038/s41591-019-0508-1

16. Wang Z, Wang P, Liu K, Wang P, Fu Y, Lu C-T, Aggarwal CC, Pei J, Zhou Y. A Comprehensive Survey on Data Augmentation. arXiv preprint arXiv:240509591. 2024.

17. Redmon J. You only look once: Unified, real-time object detection. Paper/Poster presented at: Proceedings of the IEEE conference on computer vision and pattern recognition; 2016;

18. Snoek J, Larochelle H, Adams RP. Practical bayesian optimization of machine learning algorithms. Advances in neural information processing systems. 2012;25.

19. Mooney CZ, Duval RD, Duvall R. Bootstrapping: A nonparametric approach to statistical inference. sage; 1993.

20. Paszke A, Gross S, Massa F, Lerer A, Bradbury J, Chanan G, Killeen T, Lin Z, Gimelshein N, Antiga L. Pytorch: An imperative style, high-performance deep learning library. Advances in neural information processing systems. 2019;32.

21. Brociek E, Tyminska A, Giordani AS, Caforio ALP, Wojnicz R, Grabowski M, Ozieranski K. Myocarditis: Etiology, Pathogenesis, and Their Implications in Clinical Practice. Biology (Basel). 2023;12. doi: 10.3390/biology12060874

22. Mienye ID, Swart TG, Obaido G. Recurrent neural networks: A comprehensive review of architectures, variants, and applications. Information. 2024;15:517.

